# Nomogram-based survival predictions of patients with soft tissue cancer including heart in the United States

**DOI:** 10.1101/2022.12.31.22284079

**Authors:** Roungu Ahmmad

**Affiliations:** University of Mississippi Medical Center, Department of Data Sciences

**Keywords:** Soft Tissue Sarcoma, Cancer Stages, Survival, Cancer Treatment, Stratified Cox Model

## Abstract

**Introduction:** This study aimed to assess the effects of cancer treatment on sarcoma types and to predict overall survival probabilities using nomograms.

**Methods:** This study uses the SEER-18 database, Version 2020, sponsored by the National Cancer Institute (NCI). The study cohort included participants diagnosed between 2000 and 2018 with soft tissue cancers including heart. A multivariable Cox proportional hazards model was applied to predict mortality rate, and nomograms were used to predict overall survival probability.

**Results:** The median survival time for 24,849 study participants was 48 months (IQR: 19-102) with Spindle Cell Sarcoma (ScS) having a shorter median survival time compared with Liposarcoma (LiS). A significant number of STS patients had surgery, where surgery on ScS improved survival by 75% (HR: 0.25, 95%CI: 0.19-0.32, p<0.001) and those who received radiation had a 26% improvement (HR: 0.74, 95%CI: 0.61-0.89, p=0.001). Furthermore, chemotherapy on GcS resulted in a 40% reduction in mortality for patients compared to those who did not receive chemotherapy (HR 0.60, 95%CI:0.45-0.80, p<0.001). Based on nomogram, after two, five, and ten years, a patient who received surgery on their primary sites would have a survival rate of approximately 90 percent. In contrast, a patient who did not receive surgery on their primary sites would only live for 20 percent or less. Patients with MyS have a 90% chance of surviving for 5 and 10 years after surgery, but only 22% and 10% without surgery.

**Conclusions:** Based on the results of this study, surgical and radiation intervention on sarcomas was associated with improved survival in patients with STS, while chemotherapy and primary systemic therapy had contradictory effects.

## BACKGROUND OF STUDY

Soft-tissue sarcomas (STS) are rare and heterogeneous types of malignant cells, which arise in connective tissues and other mesenchymal tissues (Demetri GD, et al., 2010) To detect disease extension, it is important to perform a correct histological diagnosis, including immunohistochemistry. Based on the molecular pathogenesis of these tumors over the past decade, there are more than 35 histologic subtypes. A survey from 2016-2018 found that 0.4 percent of males and females will be diagnosed with STS cancer during their lifetime (Saad AM, et al., 2017; Funato K, et al., 2021). According to SEER, the incidence of STS cancer is on the rise every year, and they are more likely to die from this disease (Ill EC, et al., 1996; Rouhani P, et al., 2008; Stoltzfus KC, 2020). In the United States, soft-tissue sarcoma accounts for approximately one percent of all incident cancers (Zahm, et al., 1997). Approximately 13,040 new cases of STS were estimated in 2018, with half of these deaths caused by this disease (Gutierrez JC, et al., 2007; Labrot, E., 2018)

STS were more common among patients 65 years and older (11.3 per 100,000), compared to 2.3 per 100,000 for those under 65 years of age (Miller KD, 2018; Schöffski P, et al., 2014). STS are relatively rare in the general population, but because they are typically high-grade diseases and diagnosed at an advanced stage, their survival rates are poor (Cella DF, et al., 1990). Patients with STS would have also been able to choose from a few basic treatment options, such as chemotherapy, radiation therapy, and surgery on a prime site (Fuller CD, et al., 2007). There is, however, little information pertaining to tumor histology and cancer stage’s influence on treatment outcomes, especially for elderly patients (Wright CM, et al., 2016).

Several studies have demonstrated that radiation, chemotherapy, and surgical intervention decrease the mortality rate of STS patients (Loap P, et al., 2021; Parikh RC, et al., 2018; Hoven-Gondrie ML, et al., 2016). Despite advances in surgical and radiation techniques, the prognosis for malignant pleura mausoleum patients has not improved over the past four decades. Surgery-based treatment may be beneficial for these patients, since cancer-directed surgery independently predicts improved survival and detoxed and gemcitabine were the most used chemotherapy drugs in STS patients who were older (Parikh RC, et al., 2018; Taioli E, et al., 2015). Although prognosis varies according to histological classification, older adults who develop STS with advanced stage have a poor prognosis (Sultan I, et al., 2010; Smartt AA, et al., 2020). Sarcoma types do appear to have significant genetic components and there is evidence that environmental factors may also play a role in the outcome (Warren J, et al., 2002 ; Nattinger A, 1997). A nomogram is a model-based prediction of survival probabilities that incorporates data relating to the effects of different types of treatments on different types of cancer (Nattinger A, et al., 2018; Necchi, A., 2017; Kim, S. Y., et al., 2018). Medical, oncology, and other fields have produced nomograms and projected prognoses for predicting survival rates (Qian, X., et al., 2021). Nomograms are used to determine the numerical probability of an event based on multiple prognostic and determinant variables. In these circumstances, the cancer treatment paradigms will have to be supported by nomograms, which will offer more user-friendly digital interfaces and provide more accurate prognoses.

Studying new cases, deaths, and survival rates over time (trends) enables us to determine how much progress has been made and what other research is required to address the challenges faced by patients with STS. Therefore, this study aimed to determine the mortality rate and to predict the overall survival probability for different cancer treatments and sarcoma types for patients with STS including heart cancer in the United States.

## MATERIALS AND METHODS

### Data Sources

The SEER Program is a population-based cancer database that geographically encompasses more than a quarter of the US population within its catchment areas (Hankey BF, et al., 1999). SEER program registries collect data on patient demographics, cancer type, stages, first course of treatment, and vital status (Hankey BF, et al., 1999; Frey CM, et al., 1992 ; Hankey BF, et al., 1999; Gamboa AC, et al., 2021). The study examined SEER Research Plus from 2000 through 2018 dataset 18 Registries case listings and collected data from 147 sub-setting cases. We had 7.75 million cancer cases in our case study, of which 55,261 were soft tissue cancers including heart cancer patients [Figure 1]. The most common sarcomas selected for this study were 4.5 Liposarcoma (LiS), 4.7 Leiomyosarcoma (LeS), 4.6 Synovial Sarcomas (SyS), 4.10 Spindle Cell Sarcomas (ScS), 4.14 Giant Cell Sarcomas (GcS) and 4.4.1 Myxofibrosarcomas (MyS). The dataset contained a wide variety of information regarding rare disease sarcomas, including demographic characteristics, tumor characteristics, and relevant cancer treatment information, which offered an opportunity to investigate the survival of these cancer patients. The primary objective of this study was to establish overall survival and mortality prediction. Therefore the survival time point was defined as the time from diagnosis to death and this study considered from 19 years follow up from 2000-2018. The exposure variables included whether patients received radiation therapy, chemotherapy, primary systemic treatment, and surgery on the prime site as well as baseline covariates such as age, gender, race, tumor grade, cancer stage, and total tumors in the primary site.

**Figure 1:**
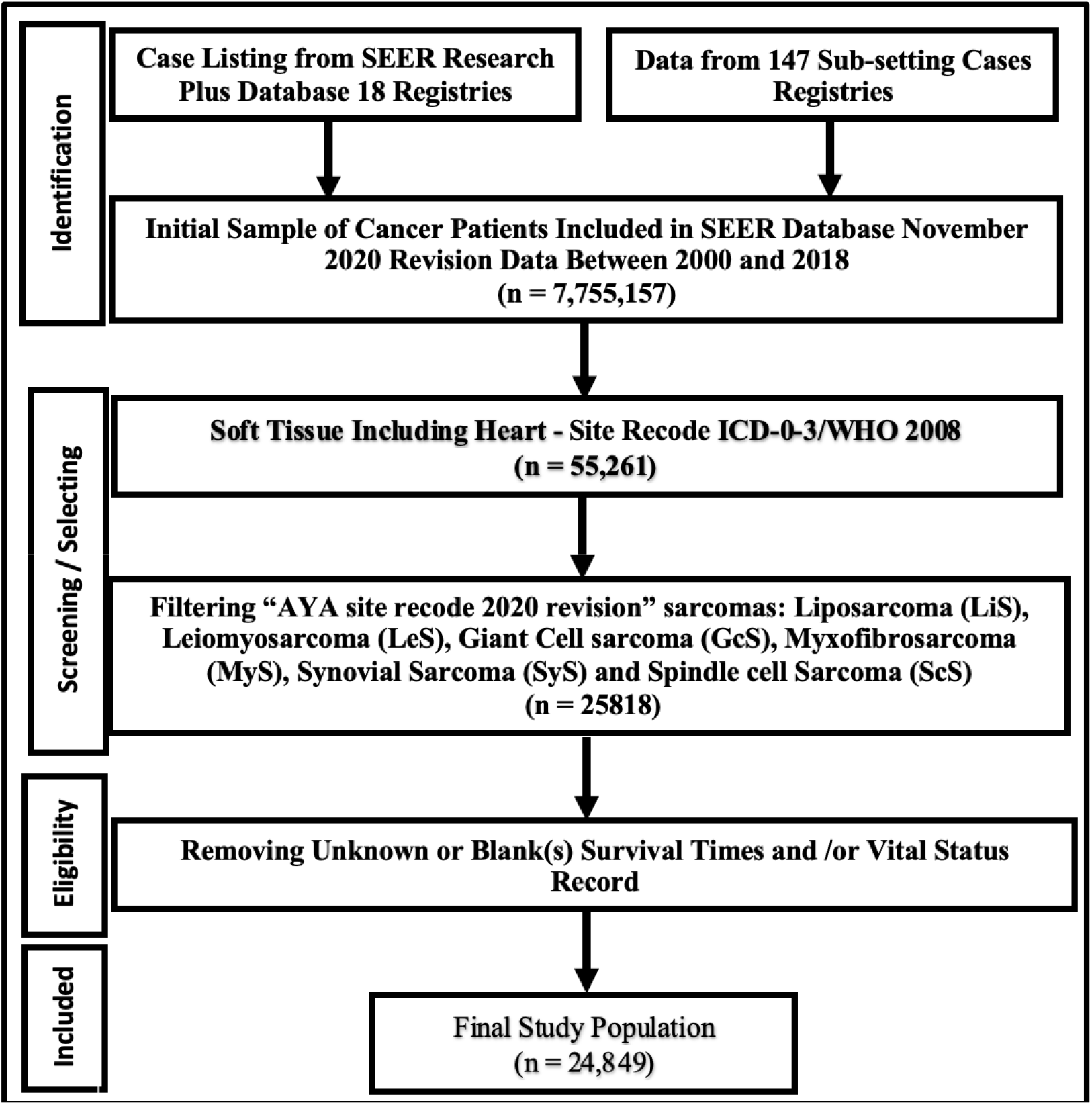
Flowchart of soft tissue including heart cancer identification in the SEER (18 registries) database. Data identification screening, eligibility and final included sample selection steps including the remove samples.

### Statistical Analysis

Data were analyzed with R statistical software and STATA version Stata/IC 15.1 for Mac (64-bit Intel) Revision 03 Feb 2020 Copyright 1985-2017 StataCorp LLC, Single-user Stata perpetual license, Serial Number 301506386406, Licensed to Md Roungu Ahmmad, The University of Mississippi Medical Center. To determine whether there were significant differences between sarcomas and covariates of interest, the Pearson Chi-square test was used. Throughout this period, the observed survival rate refers to the likelihood of survival from all causes of mortality. The Kaplan-Meier method was applied to calculate the survival rate among selected sarcomas among STS patients. We used the log rank test in order to determine whether differences in survival curves were statistically significant. All the results were considered significant level when p less than 0.05. Hazard ratios (HRs) and 95% confidence intervals (CIs) were calculated based on multivariable Cox proportional hazard models. To estimate the hazard rate for different types of sarcomas, models were developed for subsamples of patients based on the types of sarcomas. Finally developed the nomogram based on all-cause mortality for patients who received only specific cancer treatments to determine the overall survival for patients with various types of sarcomas.

## RESULTS

### Patients Characteristics

The study included 24,849 patients with STS that met the inclusion criteria (Fig. 1, Tab. 1). On average, STS patients were 59 years old at the time of diagnosis and 13,880 (56%) were males, The majority were white (n = 20,261, 82%) (Table 1). On average, patients with GcS have a considerably shorter median survival time (42 months, p<0.001), compared to other sarcomas, but those with LiS have significantly longer survival times (76 months, p<0.001). Among the selected sarcomas, LiS was the most prevalent histologic factor (n = 9098), while ScS was the least common. At the time of diagnosis, 2,775 (12%) of the patients had metastatic (distant) cancer stages, and 18,021 (73%) of the patients had at least one tumor in the primary site. Radiation therapy was administered to 42% of patients, surgery to 82%, chemotherapy to 19%, and primary systemic therapy to only 14%.

**Table 1:** Population characteristics by soft-tissue cancer types.

| Predictors | Overall,<br>N = 24,849<sup>1</sup> | LiS,<br>N = 9,098<sup>1</sup> | GcS,<br>N = 3,042<sup>1</sup> | LeS,<br>N = 6,168<sup>1</sup> | MyS,<br>N = 2,405<sup>1</sup> | ScS,<br>N = 1,880<sup>1</sup> | SyS,<br>N = 2,256<sup>1</sup> | p-value<sup>2</sup> |
| --- | --- | --- | --- | --- | --- | --- | --- | --- |
| **Time** | 65 (58) | 76 (60) | 42 (42) | 62 (58) | 71 (54) | 44 (52) | 74 (63) | <0.001 |
| **VitalStatus** |  |  |  |  |  |  |  | <0.001 |
| Alive | 13,851 (56%) | 5,912 (65%) | 1,466 (48%) | 2,788 (45%) | 1,692 (70%) | 695 (37%) | 1,298 (58%) |  |
| Dead | 10,998 (44%) | 3,186 (35%) | 1,576 (52%) | 3,380 (55%) | 713 (30%) | 1,185 (63%) | 958 (42%) |  |
| **Age** | 59 (18) | 59 (17) | 66 (16) | 62 (16) | 60 (19) | 61 (20) | 39 (19) | <0.001 |
| **Male** | 13,880 (56%) | 5,509 (61%) | 1,792 (59%) | 3,075 (50%) | 1,330 (55%) | 998 (53%) | 1,176 (52%) | <0.001 |
| **Race: White** | 20,261 (82%) | 7,427 (82%) | 2,526 (84%) | 5,048 (83%) | 1,968 (83%) | 1,491 (80%) | 1,801 (81%) | 0.002 |
| (Missing) | 259 | 88 | 34 | 63 | 36 | 17 | 21 |  |
| **Grade** |  |  |  |  |  |  |  | <0.001 |
| I | 5,091 (27%) | 4,106 (53%) | 8 (0.3%) | 541 (13%) | 328 (16%) | 84 (6.3%) | 24 (1.8%) |  |
| II | 3,862 (20%) | 1,221 (16%) | 87 (3.4%) | 1,159 (28%) | 814 (39%) | 275 (21%) | 306 (23%) |  |
| III | 3,983 (21%) | 1,100 (14%) | 515 (20%) | 1,042 (26%) | 372 (18%) | 361 (27%) | 593 (45%) |  |
| IV | 6,177 (32%) | 1,259 (16%) | 1,981 (76%) | 1,332 (33%) | 600 (28%) | 608 (46%) | 397 (30%) |  |
| (Missing) | 5,736 | 1,412 | 451 | 2,094 | 291 | 552 | 936 |  |
| **Cancer Stage** |  |  |  |  |  |  |  | <0.001 |
| Localized | 15,797 (67%) | 6,388 (74%) | 1,870 (65%) | 3,519 (61%) | 1,826 (79%) | 896 (52%) | 1,298 (60%) |  |
| Regional | 4,956 (21%) | 1,808 (21%) | 650 (23%) | 1,140 (20%) | 410 (18%) | 433 (25%) | 515 (24%) |  |
| Distant | 2,775 (12%) | 470 (5.4%) | 357 (12%) | 1,116 (19%) | 87 (3.7%) | 410 (24%) | 335 (16%) |  |
| (Missing) | 1,321 | 432 | 165 | 393 | 82 | 141 | 108 |  |
| **Total Tumor** |  |  |  |  |  |  |  | <0.001 |
| One | 18,021 (73%) | 6,592 (72%) | 2,063 (68%) | 4,398 (71%) | 1,727 (72%) | 1,292 (69%) | 1,949 (86%) |  |
| 2-3 Nodes | 6,350 (26%) | 2,330 (26%) | 912 (30%) | 1,640 (27%) | 628 (26%) | 548 (29%) | 292 (13%) |  |
| More than 3 | 470 (1.9%) | 171 (1.9%) | 66 (2.2%) | 129 (2.1%) | 50 (2.1%) | 40 (2.1%) | 14 (0.6%) |  |
| (Missing) | 8 | 5 | 1 | 1 | 0 | 0 | 1 |  |
| **Radiotherapy** | 10,397 (42%) | 3,429 (38%) | 1,718 (56%) | 1,928 (31%) | 1,185 (49%) | 827 (44%) | 1,310 (58%) | <0.001 |
| **Chemotherapy** | 4,730 (19%) | 963 (11%) | 697 (23%) | 1,282 (21%) | 203 (8.4%) | 495 (26%) | 1,090 (48%) | <0.001 |
| **SystemicTherapy** | 2,551 (14%) | 543 (8.3%) | 479 (18%) | 586 (14%) | 146 (7.4%) | 215 (16%) | 582 (39%) | <0.001 |
| (Missing) | 6,568 | 2,528 | 325 | 1,995 | 445 | 496 | 779 |  |
| **SurgeryPrimesite** | 19,855 (82%) | 7,732 (87%) | 2,429 (81%) | 4,528 (76%) | 2,218 (93%) | 1,133 (62%) | 1,815 (82%) | <0.001 |
| (Missing) | 557 | 193 | 50 | 206 | 20 | 50 | 38 |  |

<sup>1</sup> Median (IQR) or Non-Frequency (%)<sup>2</sup> Kruskal-Wallis rank sum test; Pearson's Chi-squared test

### Survival Analysis

In terms of overall survival, there was significant difference between sarcoma types for patients with STS (Fig 2). In general, survival rates for MyS patients were higher than those for all other sarcomas, while survival rates for ScS patients were lower. In patients with MyS, the overall survival rate was 75%, whereas in patients with ScS it was 37%. Furthermore, patients with LiS had a higher survival rate than those with GcS, LeS, ScS and SyS. Survival rates vary with sarcomas according to the type of cancer, its stage, how advanced it is, and other factors. Radiation therapy, chemotherapy, and surgery significantly improved overall survival among patients with STS (Fig). The factors that negatively affected survival on a multivariate analysis were increasing age, male sex, non-whites, and advanced cancer stages (Fig. 3). In the present study, ScS, LiS, GcS, LeS, and MyS all had significant associations with patient survival, where patient with ScS had a higher likelihood of dying whereas MyS patients had a lower likelihood (Fig 3). As compared with untreated patients, radiotherapy, chemotherapy, and surgery significantly reduced the mortality rate, although systemic therapy provided a contrary outcome by adjusting all sarcomas. In metastasis stage cancer patients had 3.66-fold higher hazard of death compare with patients in primary stages (HR 3.66, 95% CI: 3.34 - 4.00, p<0.001). In this manner, the tumor grade and stage significantly affect patient survival. As the tumor grade and stage increase, the mortality rate for patients also increases. Detailed analysis of survival among sarcoma patients was conducted using subsample Cox proportional hazard model by adjusting for cancer staging, tumor histological features, and demographic factors. (Tab 2).

**Figure 2:**
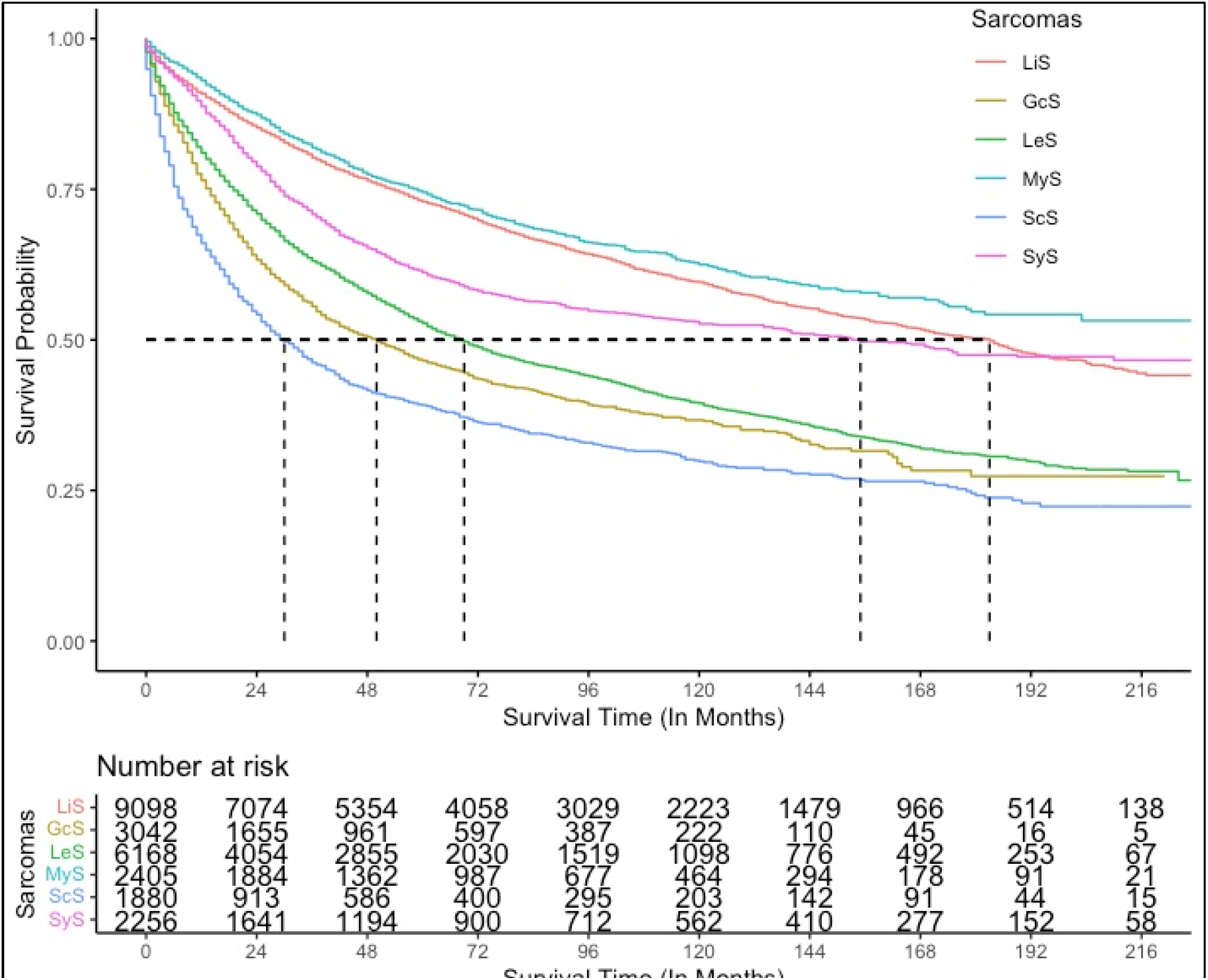
The figure shows stratified Kaplan-Meier survival estimates for soft tissue sarcom

**Figure 3:**
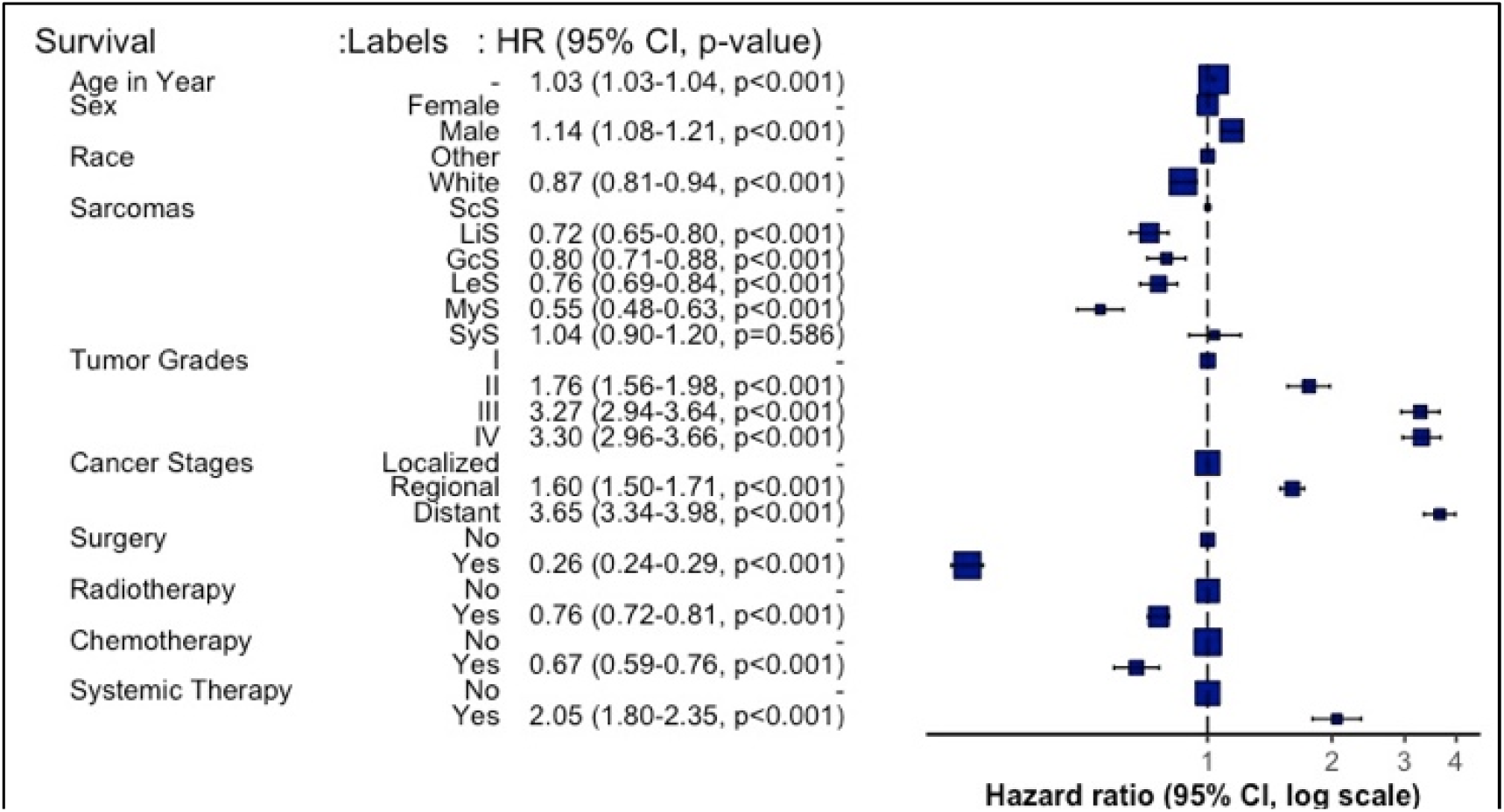
Overall Hazard and 95% Confidence interval including hazard forest

Table 2 presents the results of the Cox proportional hazards model for sarcomas. For every additional year of patient age on diagnosis, the risk of death increased by 2%, 5%, 3%, and 6% for ScS (HR: 1.02 95%CI: 1.02-1.03), LiS (HR:1.05, 95%CI: 1.04-1.05), GcS (HR: 1.03, 95%CI: 1.02-1.03) and MyS (HR:1.06, 95%CI: 1.05-1.07) respectively. The mortality rate of patients with various sarcomas who underwent surgery at the prime site was significantly reduced in comparison to those who did not undergo surgery. Patients with MyS STS who underwent surgery had a reduced mortality rate of 71% (HR 0.29, 95% CI: 0.19-0.43) in comparison with those who did not. Similar findings were reported for ScS, LiS, GcS LeS, and SyS that showed a substantial reduction in mortality among patients who received surgical intervention. For patients with sarcomas other than MyS, chemotherapy and radiation treatment significantly reduced mortality. However, primary systemic treatment had a contradictory effect.

**Table 2:**
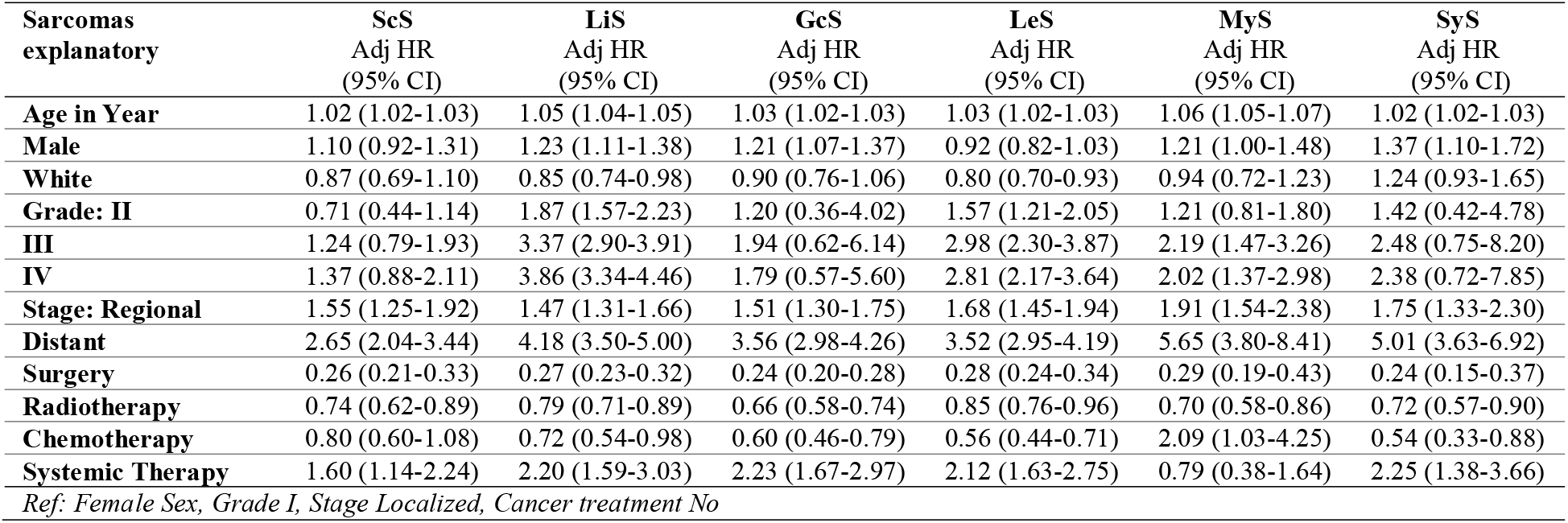
Hazard Ratios and 95% Confidence Interval for Multivariate Cox model of Overall Mortality, stratified by cancer treatment with adjusting demographic and tumor characteristics factors

| Sarcomas explanatory | ScS<br>Adj HR<br>(95% CI) | LiS<br>Adj HR<br>(95% CI) | GcS<br>Adj HR<br>(95% CI) | LeS<br>Adj HR<br>(95% CI) | MyS<br>Adj HR<br>(95% CI) | SyS<br>Adj HR<br>(95% CI) |
| --- | --- | --- | --- | --- | --- | --- |
| Age in Year | 1.02 (1.02-1.03) | 1.05 (1.04-1.05) | 1.03 (1.02-1.03) | 1.03 (1.02-1.03) | 1.06 (1.05-1.07) | 1.02 (1.02-1.03) |
| Male | 1.10 (0.92-1.31) | 1.23 (1.11-1.38) | 1.21 (1.07-1.37) | 0.92 (0.82-1.03) | 1.21 (1.00-1.48) | 1.37 (1.10-1.72) |
| White | 0.87 (0.69-1.10) | 0.85 (0.74-0.98) | 0.90 (0.76-1.06) | 0.80 (0.70-0.93) | 0.94 (0.72-1.23) | 1.24 (0.93-1.65) |
| Grade: II | 0.71 (0.44-1.14) | 1.87 (1.57-2.23) | 1.20 (0.36-4.02) | 1.57 (1.21-2.05) | 1.21 (0.81-1.80) | 1.42 (0.42-4.78) |
| III | 1.24 (0.79-1.93) | 3.37 (2.90-3.91) | 1.94 (0.62-6.14) | 2.98 (2.30-3.87) | 2.19 (1.47-3.26) | 2.48 (0.75-8.20) |
| IV | 1.37 (0.88-2.11) | 3.86 (3.34-4.46) | 1.79 (0.57-5.60) | 2.81 (2.17-3.64) | 2.02 (1.37-2.98) | 2.38 (0.72-7.85) |
| Stage: Regional | 1.55 (1.25-1.92) | 1.47 (1.31-1.66) | 1.51 (1.30-1.75) | 1.68 (1.45-1.94) | 1.91 (1.54-2.38) | 1.75 (1.33-2.30) |
| Distant | 2.65 (2.04-3.44) | 4.18 (3.50-5.00) | 3.56 (2.98-4.26) | 3.52 (2.95-4.19) | 5.65 (3.80-8.41) | 5.01 (3.63-6.92) |
| Surgery | 0.26 (0.21-0.33) | 0.27 (0.23-0.32) | 0.24 (0.20-0.28) | 0.28 (0.24-0.34) | 0.29 (0.19-0.43) | 0.24 (0.15-0.37) |
| Radiotherapy | 0.74 (0.62-0.89) | 0.79 (0.71-0.89) | 0.66 (0.58-0.74) | 0.85 (0.76-0.96) | 0.70 (0.58-0.86) | 0.72 (0.57-0.90) |
| Chemotherapy | 0.80 (0.60-1.08) | 0.72 (0.54-0.98) | 0.60 (0.46-0.79) | 0.56 (0.44-0.71) | 2.09 (1.03-4.25) | 0.54 (0.33-0.88) |
| Systemic Therapy | 1.60 (1.14-2.24) | 2.20 (1.59-3.03) | 2.23 (1.67-2.97) | 2.12 (1.63-2.75) | 0.79 (0.38-1.64) | 2.25 (1.38-3.66) |

Ref: Female Sex, Grade I, Stage Localized, Cancer treatment No

## CLINICAL PERSPECTIVES OF STUDY

Nomograms can guide the treatment for different types of sarcomas and improve survival among patients with STS. A number of studies have revealed nomograms and estimated prognoses in oncology and medicine [1,4]. By integrating diverse prognostic and determinant factors, nomograms can calculate a numerical probability of a clinical outcome for individualized medicine [2,5]. An all-causes mortality nomogram was used to estimate survival probability for sarcomas based on the adjustment of baseline predictors and tumor histological factors. In FIG 4, overall survival probability was predicted using all-causes mortality nomograms by adjusting patient’s age, sex race, cancer stage, grade, and cancer treatments.

**Figure 4:**
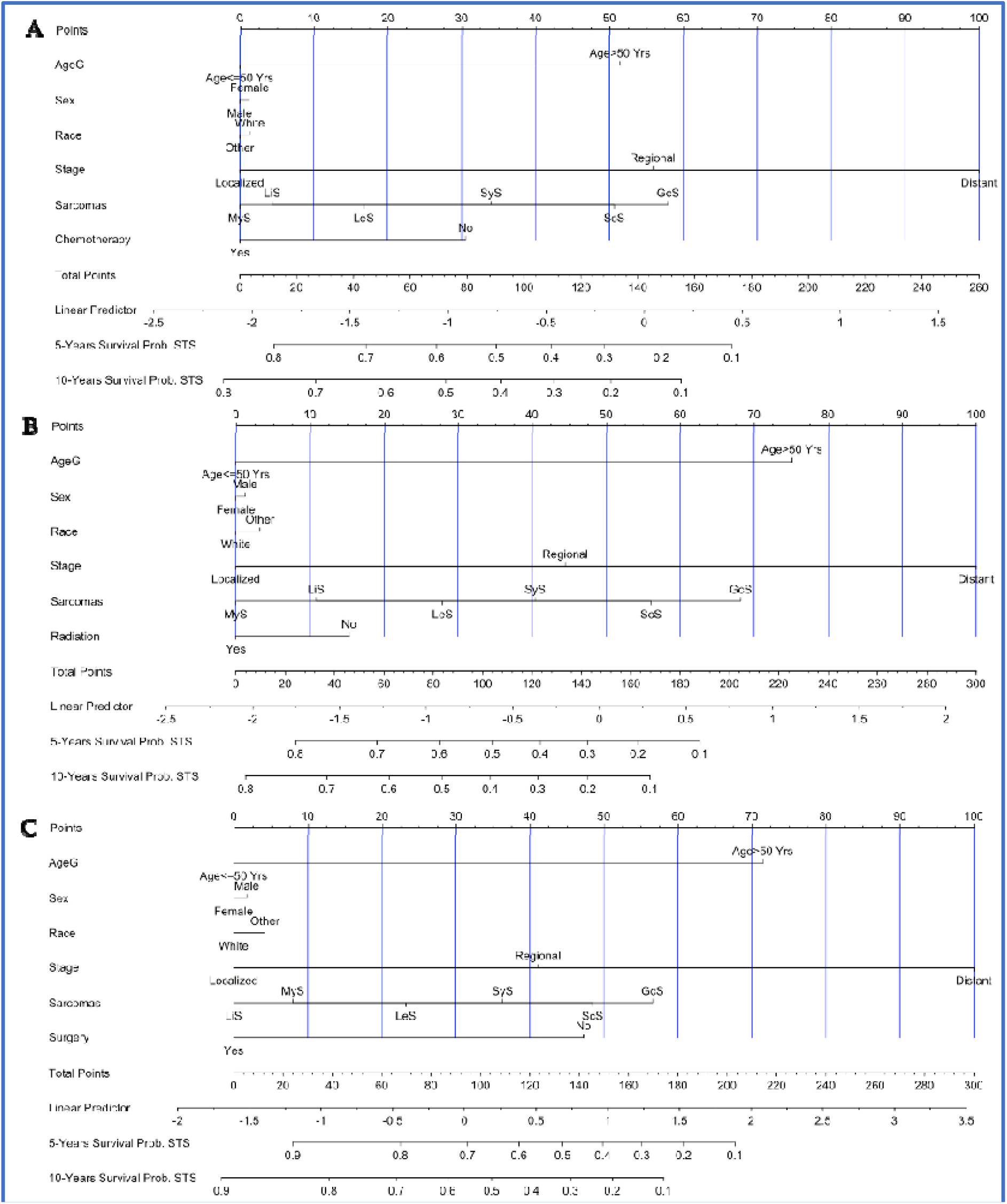
Overall survival nomogram and nomogram only sarcomas and treatment combination. This nomogram provides a method to calculate 5 and 10-year probability of survival based on a patient’s combination of covariates. Draw a line straight down to the 5-year and 10-year overall survival axes to obtain the probability. A: The patient who received only Chemotherapy; B: The patients who received only Radiotherapy and C: The patient who received only Surgical Intervention.

Assigning a point value to each variable is accomplished by first locating the patient’s characteristics row and then drawing a vertical line straight up to the points row. By drawing a vertical line directly up to the point row in Fig 4A, for example, we obtain a score of 32 points for the patient who did not receive chemotherapy. Repeat this process for each row, accumulating points for each variable. The overall survival probability is determined by adding up all the points and drawing a vertical line from the total points column. The overall survival probability for patients who did not undergo chemotherapy at the end of 5 years would be 55%, and at the end of 10 years it would be 46%. On the other hand, for patients who underwent chemotherapy, the overall survival rate after 5 and 10 years would be greater than 80% (Figure 4A). Moreover, patients who had only surgery on the primary sites would survive 5 and 10 years approximately above 90%, while those who did not receive surgical intervention would only survive 45% and 25% respectively (Figure 4C). Patients with STS who received only radiation therapy did not improve their survival significantly (Figure 4B) in this study. Considering all cancer treatments, chemotherapy, radiation therapy, and surgery have an 85% improvement in survival rate for patients with MyS. However, for patients with GcS, five-year survival rates are very low (18% for chemotherapy, 10% for radiation therapy, and 25% for surgery). In all the nomograms, calibration was very closely matched between the actual and predicted event probability in the test dataset, and the internal and external calibration of the all-causes mortality nomogram showed an excellent correlation between prediction and observation.

## DISCUSSION

The purpose of the study was to investigate the impact of cancer treatment on patients with soft tissue including heart cancer as part of a large population-based cohort study (SEER). This is the first study to investigate cancer treatments for various types of sarcomas by assessing tumor histological factors, cancer stages, and developing a reliable model-based nomogram prediction. Our findings show that the factors negatively affecting on survival were increasing age, male sex, non-whites, increasing cancer stages, and tumor grade. Additionally, receiving cancer treatments (surgery, chemo, and radiotherapy) significantly reduced the probability of death for patients with STS, including heart cancer.

Several studies have demonstrated increasing ages and advanced cancer stages for patient with STS, the mortality rate significantly increased (Gutierrez, J. C., et al., 2007; Ognjanovic, S., et al., 2009; Gazendam, A. M., et al., 2021). In general, patients at an advanced stage, those who do not receive cancer treatment, and those who are older have higher mortality rate. Approximately sixty seven percent of patients were diagnosed at an earlier stage in this study and progressed to STS with a mean progression time of 65 months. There are 12% of patients whose disease is in distant stages, and they represent 1,116 (19%) of the LeS cancer types. Consistent with previous studies, LeS was the most common histological category of STS in the study population. Among patients with STS, there was a significant difference in survival between types of sarcomas. MyS patients had a higher survival rate than patients with other sarcomas, while those with ScS had a lower survival rate. It has been noted that the overall survival rate of patients with MyS is 75%, whereas the overall survival rate of patients with ScS is 37%. In addition, patients with LiS have a higher survival rate than those with GcS, LeS, ScS, and SyS. Based on previous studies, ScS is the second most common sarcoma and has a lower survival rate, which is consistent with our findings (Ramlawi, B., et al., 2016; Agaram, N. P., 2019).

Most patients in this study were older adults who underwent surgery on the prime site (83%) and radiation therapy (42%). This study observed a relatively lower proportion of patients treated with chemotherapy (19%) and primary systemic therapy (14%). In this study, we try to shed some light on some of the related issues with sarcomas in older patients. A lower mortality rate was observed for STS patients who underwent chemotherapy, radiation, and surgery at the primary site, in line with other studies (Gross, J. L., et al., 2005; Pasquali, S., et al., 2019; Woll, P. J., et al., 2012). A patient with cancer of the soft tissue could receive chemotherapy, surgery or radiation therapy at the primary site. There may be different combinations of treatment that might result in different survival rates. According to some studies, patients with soft tissue cancer may have a worse prognosis after undergoing treatment (Götzl, R., et al., 2019). However, the question of which treatment would mean a higher chance of survival remains very controversial. Several studies have employed the Cox model to estimate overall survival of STS patients based on SEER data for different time points and objectives (Amer, K. M.,, et al., 2019; Wang, X.,et al., 2020). The study found that surgery at the primary site significantly reduced mortality by 74%, and chemotherapy and radiation significantly reduced mortality by 33% and 24%, respectively. Similar results were observed for patients with SyS, ScS, MyS, LeS, and LiS. Compared to radiation or chemotherapy, surgery significantly reduced mortality rates. Several studies have shown that radiation and chemotherapy can be analogous to each other in treating sarcoma, since both treatments offer different degrees of radical mastectomy or amputation, and chemotherapy offers different benefits for local control of cancer types (Dirksen, U., et al., 2008; Mortezaee, K., et al., 2019).

Furthermore, we analyzed all-cause mortality nomograms based on multivariate Cox models for STS patients. In several studies, survival has been predicted by nomograms for breast cancer, prostate cancer, melanoma, thyroid cancer, and head and neck squamous cell carcinoma by different study times and endpoints (Zhang, Y., et al., 2019; Zhou, Z., et al., 2019; Yang, K.,; Botticelli, A., et al., 2019). In this study, we are the first to develop self-evaluation algorithms for the prediction of survival probability for patients with STS including heart cancer. First, we selected patients who had only received chemotherapy, and then we fitted the multivariate Cox model to determine the hazard rates. Following that, we built a predicted nomogram in order to predict the overall survival probability of the patient.

A similar process was applied to other cancer treatments. At the end of five years and ten years, patients who did not receive chemotherapy would have a survival probability of 55% and 46%, respectively. After five or ten years, chemotherapy patients would have an overall survival rate greater than 80 percent. Moreover, patients who had only surgery on the primary sites would survive 5 and 10 years approximately above 90%, while those who did not receive surgical intervention would only survive 45% and 25% respectively. Patients with STS who received only radiation therapy did not improve their survival significantly. For patients with MyS, cancer treatments have an 85% increase in survival rates. In contrast to that, the five-year survivability rates of patients with GcS are relatively poor (18% for chemotherapy, 10% for radiation therapy, and 25% for surgery). In all the nomograms, calibration was very closely matched between the actual and predicted event probability in the test dataset, and the internal and external calibration of the all-causes mortality nomogram showed an excellent correlation between prediction and observation.

## LIMITATIONS

Despite the fact that we used a large dataset from SEER, our study had some limitations. Some clinicopathological factors (such as surgical margins, perineural invasion, solid tumors, and p53 positivity) have not been examined due to the lack of public access to the SEER dataset. In addition, although histological details have been found to be crucial to survival prediction, we cannot include them in the model because no histological subtypes of STS are included in the SEER public-use dataset. In addition, SEER does not provide information regarding recurrence of cancer. Due to a lack of information on radiotherapy techniques (total dose, fraction size, beam energy), results could not be used to determine whether survival varies with these factors. In addition, this study has adjusted for all patient and tumor characteristics despite the fact that some confounding variables were not available in this dataset (for example, tumor depth, margin, and performance status). In cases of low-grade sarcomas, adjuvant radiation may be indicated if certain risk factors (e.g., incomplete resection, positive margins) are not found in the SEER database. Another drawback of this study is the short duration of follow-up, since only a few patients were available during the study period. A longer follow-up could improve the accuracy of the model, particularly in patients suffering from STS. It is possible that the nomogram and the model perform well, but external validation of its effectiveness with other cohorts is still required.

## CONCLUSIONS

The study was performed by one of the largest population-based surveys of cancer patient which included soft tissue including heart in the United States. The factors that negatively affected on survival were increasing age, male sex, non-whites, and advanced cancer stages. Sarcomas associated with MyS have higher survival rates, but those associated with ScS have worsened survival. A better survival outcome was associated with surgery, radiation therapy and chemotherapy when treating patients with STS, but primary systemic therapy had conflicting results. An all-cause mortality nomogram was developed and predicts overall survival probability of STS patients.

## Data Availability

all meta data will be provided upon request

## ACKNOWLEDGMENTS

The authors would like to thank the NCI for open access to their SEER database. The opinions or views expressed in this paper are those of the authors and do not represent the opinions or recommendations of the NCI.

## FUNDING SOURCES

No grant funding was received for this study

## DISCLAIMER

The views expressed in this manuscript are those of the authors and do not necessarily represent the views of the National Cancer Institute (NCI); or the U.S. Department of Health and Human Services.

## Notes

### Competing Interest Statement

The authors have declared no competing interest.

### Funding Statement

No funding

### Author Declarations

The SEER Program is a population-based cancer database that geographically encompasses more than a quarter of the US population within its catchment areas (Hankey BF, et al., 1999). SEER program registries collect data on patient demographics, cancer type, stages, first course of treatment, and vital status (Hankey BF, et al., 1999; Frey CM, et al., 1992 ; Hankey BF, et al., 1999; Gamboa AC, et al., 2021). The study examined SEER Research Plus from 2000 through 2018 dataset 18 Registries case listings and collected data from 147 sub-setting cases

